# Modelling the tail-phase pharmacokinetics of long-acting cabotegravir and rilpivirine from early pregnancy to postpartum at steady state

**DOI:** 10.64898/2026.04.02.26350020

**Authors:** Shakir Atoyebi, Catriona Waitt, Adeniyi Olagunju

**Author notes:** **Corresponding author:** Dr Shakir Atoyebi. Department of Biochemistry, Cell, and Systems Biology, Biosciences Building, Crown Street, Liverpool, L69 7BE, United Kingdom.

## Abstract

Long-acting cabotegravir and rilpivirine combination (LA-CAB/RPV) is approved for HIV treatment whilst long-acting cabotegravir alone (LA-CAB) is approved for HIV prevention, both in adults. However, individuals who become pregnant might prefer to discontinue it due to lack of definitive data on safety. The aim of this study was to characterise the tail-phase maternal and fetal pharmacokinetics of LA-CAB/RPV following discontinuation at steady-state early in pregnancy. A virtual population of non-pregnant women (n = 100 per scenario) initiated intramuscular injections of LA-CAB/RPV at the approved dosage and continued maintenance dose (400/600 mg once monthly or 600/900 mg once every two months) until steady state. We simulated discontinuation at steady state after only one injection during pregnancy. Tail-phase pharmacokinetics of CAB and RPV from LA injections were characterised during gestation and until 6 months postpartum. Pharmacokinetic tails of LA-CAB/RPV were driven by the residual drug in the muscle depot which stabilised at steady state and reduced steadily upon dosing discontinuation. Upon discontinuation of the monthly dosing, predicted median (IQR) maternal plasma concentrations for LA-CAB were 415 (386-448) ng/mL at delivery and 125 (115-139) ng/mL 6 months postpartum. For LA RPV, these were 11.6 (11.0-12.6) ng/mL and 7.84 (7.30-8.49) ng/mL at delivery and 6 months postpartum, respectively. Pharmacokinetic tails of LA-CAB/RPV extend to several months postpartum, with levels falling below established minimum effective concentration in most women after gestation week 33. Potential strategies to minimise potential risks associated with LA-CAB/RPV discontinuation in this population are needed.

## Introduction

Over 21 million women and girls were recently estimated to be living with HIV which amounts to 53% of the global burden of HIV (1). Antiretroviral therapy has improved the life expectancy and quality of life of people living with HIV; an estimated 83% of women aged 15 years and above had access to HIV treatment in 2024 (1-6). Despite the benefits of antiretroviral therapy, some factors affecting drug adherence limit the use of oral antiretroviral drugs for optimal prevention and treatment of HIV. These factors include dosing frequency, pill burden, concerns about disclosure, and fear of stigma among others (7-9). Moreover, poor drug adherence risks virologic failure, HIV transmission and the development of drug resistance (7, 10). Long-acting injectable (LAI) antiretroviral options offer relief from daily adherence barriers but create a prolonged tail-phase after discontinuation, during which plasma concentrations decline slowly and may become subtherapeutic (11, 12). Long-acting injectable cabotegravir and rilpivirine combination (LA-CAB/RPV) is licensed as a complete regimen for the treatment of HIV-1 infection in adults who are virologically suppressed with every 4 weeks (Q4W) or every 8 weeks (Q8W) dosing after an optional oral lead-in (13, 14).

Tail-phase of long-acting drugs could be defined as the prolonged post discontinuation interval with declining, sometimes subtherapeutic plasma levels due to slow depot release and long apparent half-lives. Landovitz et al (2020) reported LA-CAB remained detectable in the body 76 weeks after dosing discontinuation in some individuals. Further analyses suggested that LA-CAB could remain detectable in the body for up to 3 years depending on sex and body mass index of the individual (12). For many anti-infective drugs, drug concentrations in the body must be kept above certain clinical thresholds to maintain effect which led to commonly used indices like multiples of the protein-adjusted drug inhibitory drug concentration. Thus, there could be an increased drug resistance risk if concentrations hover below efficacy targets for weeks to months during the tail-phase without the introduction of other suitable antiretroviral drugs.

HIV can be transmitted during pregnancy, at delivery and during breastfeeding (15, 16), but antiretroviral therapy could reduce the risk of vertical HIV transmission to < 1% (16-18). In 2024, 84% of pregnant women living with HIV had access to antiretroviral treatment to prevent vertical transmission of HIV to their child (1). However, pregnancy brings multiple physiological changes which can affect drug disposition (13, 19). Pregnancy-associated increases in UGT1A1 and CYP3A activity, expanded plasma volume, and reduced protein binding may reduce drug exposures with uncertain implications for depot driven tail concentrations of both cabotegravir and rilpivirine (20-22). Notably, reduction in rilpivirine exposure has been predicted for LA-RPV initiated during pregnancy which raises concerns about maintaining efficacy during pregnancy (23). In addition, the foetus is exposed to maternal antiretroviral drugs taken during pregnancy which potentially serves as pre-exposure prophylaxis before delivery (24).

Nonetheless, due to lack of definitive data on safety (and efficacy), some women who become pregnant may wish to discontinue LA-CAB for PrEP or LA-CAB/RPV for antiretroviral treatment during gestation (25, 26). However, LA-CAB/RPV has a long wash-out period and drug concentrations of cabotegravir and rilpivirine might persist in the body system beyond the period of gestation to lactation when stopped close to the conception period (25). Thus, though LAI administration could be stopped shortly before or early in pregnancy, the developing embryo/foetus would likely be exposed to the drug being released from the muscle depot throughout gestation. This exposure might also persist after the delivery of the baby with potential drug exposure during lactation.

Physiologically-based pharmacokinetic (PBPK) modelling is a computational tool increasingly used during drug research and development to predict drug disposition and guide related clinical studies (27). PBPK models can facilitate exploration of dose adjustments in complicated clinical scenarios like drug-drug interactions, renal and hepatic impairment (28-32). In addition, they are often employed to predict drug disposition in specific populations like pregnant women and paediatrics (33-37). More recently, we have used PBPK models to predict the disposition of long-acting antiretroviral and antipsychotic drugs during pregnancy (13, 38). In our prior modelling study, LA-CAB/RPV was initiated during pregnancy depicting clinical scenarios without earlier drug exposure before pregnancy. Here, we model discontinuation at early pregnancy after pre-conception steady state—quantifying maternal, fetal, and postpartum tail exposures by regimen. Currently, no study has quantified both the mother–fetus ‘tail-exposures’ after stopping at early pregnancy once steady state is established preconception, across Q4W and every two months regimens. Though Patel et al (2023) reported the tail exposures of LA-CAB/RPV in the mother after discontinuing, fetal exposure during this phase was not reported (25). Also, many of the participants did not reach steady state at the time of dosing discontinuation. In addition, some of the study participants were switched to drugs known to induce or inhibit enzymes that metabolise cabotegravir or rilpivirine which could have impacted their disposition during the tail phase. If drug concentrations during the tail-phase fall below minimum effective concentrations for sustained periods, the risk of virologic rebound and resistance selection would increase. Thus, subtherapeutic drug concentrations during the tail-phase could compromise virologic suppression and select drug resistance in treatment settings if oral coverage is inadequate. Prolonged low levels of rilpivirine increase the risk of drug resistance to non-reverse transcriptase inhibitors. Likewise, fetal exposure during gestation to subtherapeutic drug concentrations also poses uncertain benefit/risk.

In this study, we aimed to (1) quantify maternal tail pharmacokinetics of cabotegravir and rilpivirine after discontinuation at early pregnancy following preconception steady state, and (2) predict fetal and postpartum exposure. Clinically relevant metrics such as time to subtherapeutic levels and cord-to-maternal ratios would also be estimated to inform decisions on oral coverage and counselling.

## Materials and Methods

The development and validation of the adult and pregnancy physiologically based pharmacokinetic (PBPK) models used for this study have been previously reported (13). The model had been validated with clinical datasets of oral cabotegravir, rilpivirine and raltegravir in adults, clinical datasets of LA-CAB/RPV in adults, clinical datasets of oral rilpivirine and raltegravir during pregnancy, and clinical data of LA-CAB/RPV during wash-out after discontinuation early in pregnancy (13, 25, 39-47). The model was utilised on SimBiology (MATLAB, R2019a). This current study explored the tail-phase kinetics of LA-CAB/RPV if discontinued in early pregnancy after steady state has been reached before conception. As illustrated in Figure 1, model simulations were conducted through 3 different conditions: before, during and after pregnancy (i.e. postpartum). An adult female PBPK model was used to simulate the non-pregnant populations before pregnancy and during postpartum (Figure 1). On the other hand, two distinct pregnancy PBPK models were used for the simulations in pregnancy to account for the different phases of fetal development. A pregnancy PBPK model without a fetal sub-model was used to simulate the virtual pregnant cohort during the first trimester of pregnancy (i.e. up to week 13). In addition, a pregnancy PBPK model with a fetal sub-model was used to simulate the maternal pharmacokinetics and fetal drug exposure during the second and third trimesters of pregnancy (i.e. >week 13 – week 40).

**Figure 1:**
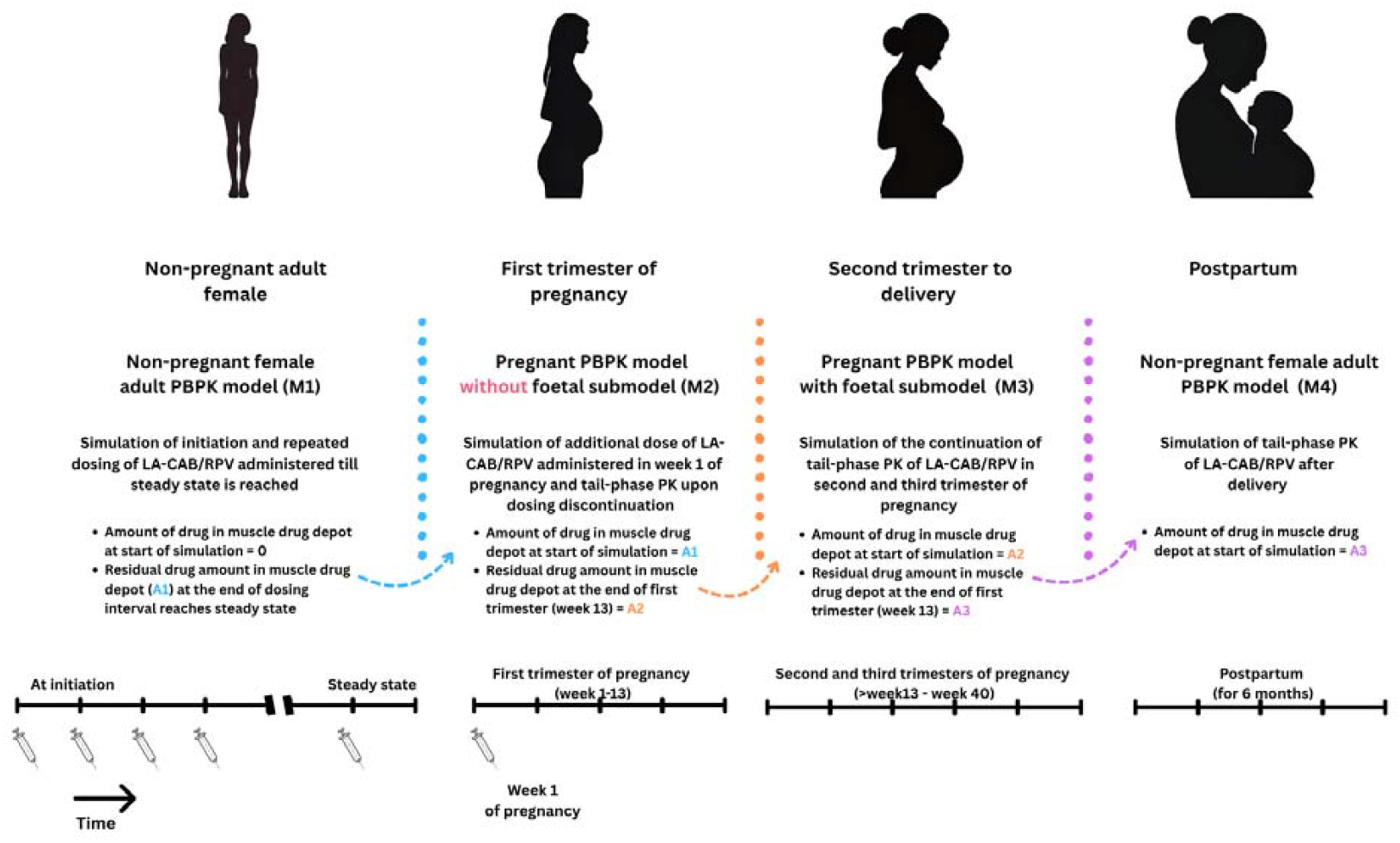
Modelling workflow for simulations for tail-phase pharmacokinetics of LA-CAB/RPV discontinued early in pregnancy when steady state is reached before conception. The figure illustrates the different populations, distinct model types used, and their related simulations.

Approved dosing of cabotegravir and rilpivirine could start with an optional 1-month daily oral lead-in with 30 mg cabotegravir, and 25 mg rilpivirine taken once daily. After the oral lead-in or without it, LA-CAB/RPV dosing starts with initial intramuscular (IM) injections of 600 mg LA-CAB and 900 mg LA-RPV at separate gluteal sites during the same visit. Thereafter, the IM dosing continues with 400/600 mg Q4W dosing of LA-CAB/RPV or Q8W dosing with 600/900 mg (48). Similarly, LA-CAB administered alone (without LA-RPV) has been approved for HIV pre-exposure prophylaxis (PrEP) with its dosing being identical to the Q8W dosing of LA-CAB when administered for treatment (49).

The dosing administration protocol employed in the model was based on the approved dosing of LA-CAB/RPV without the optional oral lead-in dose described above because reports have shown comparable drug levels with or without the oral lead-in for LA-CAB/RPV [ref]. Thus, LA-CAB/RPV, without oral lead-in, was initiated and continued to steady state in a virtual population of 100 non-pregnant women. Dosing was continued till steady state was achieved in the non-pregnant state, continued through the period of conception and terminated in early pregnancy in the virtual cohort of 100 women. The last dose was administered in week 1 of pregnancy.

Simulations of the tail-phase pharmacokinetics of LA-CAB/RPV were continued in virtual populations of pregnant women from second trimester until delivery, and from delivery until 6 months postpartum. Continuous simulation of drug disposition from pre-pregnancy to the postpartum period was enabled via transfer of the final residual drug in muscle depot to the next stage (Figure 1). Both maternal and fetal exposure measures were computed from concentration-time data for pregnancy (weeks 15-40) and postpartum. Maternal plasma concentrations during pregnancy and postpartum were compared against a value four times the protein binding-adjusted 90% inhibitory concentration (4*PA-IC_90_) for both cabotegravir and rilpivirine, which is often accepted as providing therapeutic plasma concentrations. In addition, the time for the plasma drug concentration to reach below its 4*PA-IC_90_ after the last dose was also computed.

## Results

A total of 4 scenarios were simulated (n = 100) for each LA dosing regimen in virtual cohorts of non-pregnant, pregnant and postpartum women (Figure 1). The median (interquartile range) age was 26.5 years (22.2-30.8) for all virtual cohorts. Also, the median (interquartile range) body weight for the pregnant and postpartum cohort was 64.1 kg (54.8-73.8) and 59.9 kg (51.5-68.5), respectively.

The pharmacokinetic tails for LA-CAB and LA-RPV were driven by the residual drug in the muscle depot which stabilised at steady state (Figure 2). Figure 2 shows the predicted plasma concentration profiles with the simulation of the tail-phase pharmacokinetics for the LA-CAB/RPV for both Q4W and Q8W dosing. With the Q4W dosing, the simulated residual drug in the muscle depot was 1546 mg and 2309 mg at steady state before pregnancy, 925 mg and 1383 mg at the end of the first trimester, and 208 mg and 312 mg at the end of pregnancy (week 40), for cabotegravir and rilpivirine, respectively. With the Q8W dosing, the simulated residual drug in the muscle depot was 1028 mg and 1542 mg at steady state before pregnancy, 774 mg and 1161 mg at the end of the first trimester, and 174 mg and 262 mg at the end of pregnancy, for cabotegravir and rilpivirine, respectively.

**Figure 2:**
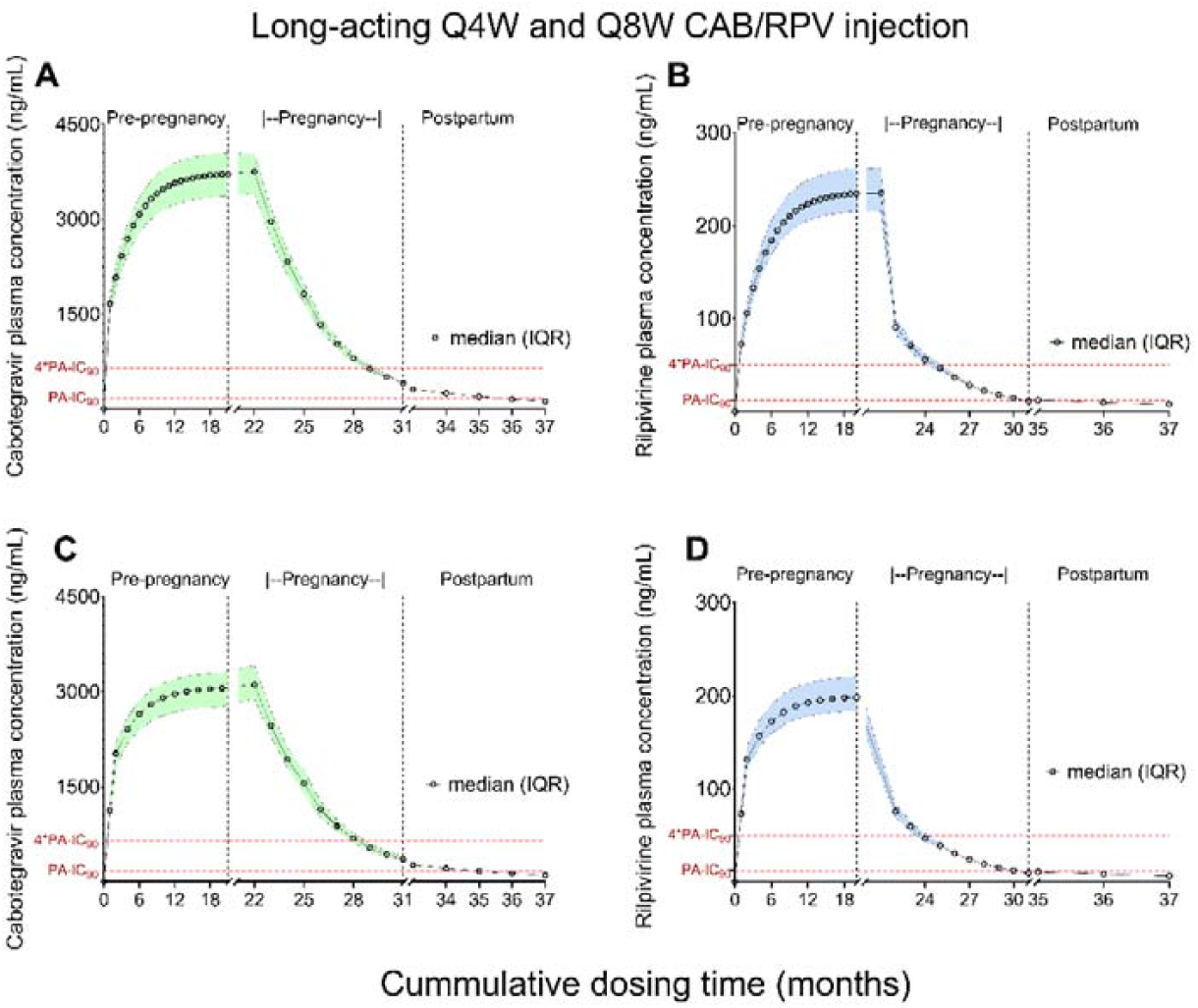
Predicted tail-phase pharmacokinetics of monthly (Q4W) and bimonthly (Q8W) long-acting cabotegravir/rilpivirine (LA-CAB/RPV) from pre-conception to 6 months postpartum. Panels show (A) Q4W cabotegravir (CAB); (B) Q4W rilpivirine (RPV); (C) Q8W cabotegravir and (D) Q8W rilpivirine total plasma concentrations (ng/mL). Curves depict the median with shaded bands indicating the interquartile range (IQR) across virtual participants (N = 100 per scenario). Vertical reference lines mark conception and delivery; background shading indicates pre-conception dosing to steady state, pregnancy tail (no further injections), and postpartum tail. Horizontal dashed lines denote protein-adjusted inhibitory thresholds: PA-IC_90_ and 4×PA-IC_90_ (CAB: 166 and 664 ng/mL; RPV: 12 and 50 ng/mL, respectively). Abbreviations: CAB, cabotegravir; RPV, rilpivirine; Q4W, every 4 weeks; IQR, interquartile range; PA-IC_90_, protein-adjusted 90% inhibitory concentration.

Maternal plasma levels were predicted to be above the respective 4*PA-IC_90_, (664 ng/mL) for cabotegravir and rilpivirine (50 ng/mL), with both drugs in 100% of the cohort at gestation week 8, compared to 0% at delivery (Table I). For LA-CAB, cabotegravir plasma concentrations were predicted to fall below the 4*PA-IC_90_ at a median (range) time of 30.1 weeks (24.4-36.1) and 27.4 weeks (13.1-32.7) after the last dose for Q4W and Q8W, respectively, during pregnancy. In addition, cabotegravir plasma concentrations were predicted to fall below the PA-IC_90_ at a median (range) time of 58.0 weeks (52.7-63) and 55.1 weeks (48.6-61.3) after the last dose for Q4W and Q8W, respectively, which occurs postpartum.

**Table 1:** Predicted proportion of women with plasma drug concentration above the minimum effective concentration.

|  | Every 4 weeks dosing |  | Every 8 weeks dosing |  |
| --- | --- | --- | --- | --- |
|  | CAB | RPV | CAB | RPV |
| **% Above 4\*PA-IC<sub>90</sub>** |  |  |  |  |
| Gestation week 8 | 100 | 100 | 100 | 98 |
| At delivery (40 weeks) | 0 | 0 | 0 | 0 |
| **% Above PA-IC<sub>90</sub>** |  |  |  |  |
| At delivery (40 weeks) | 100 | 38 | 100 | 5 |
| 5 Months postpartum | 33 | 7 | 0 | 0 |
| 6 Months postpartum | 4 | 2 | 0 | 0 |

Abbreviations: CAB – cabotegravir; GW – gestation week; MEC – minimum effective concentration; PA-IC<sub>90</sub> – protein binding-adjusted 90% inhibitory concentration (166 ng/mL for cabotegravir and 12 ng/mL for rilpivirine); RPV – rilpivirine.

With LA-RPV, rilpivirine plasma concentrations were predicted to fall below the 4*PA-IC_90_ at a median (range) time of 13.3 weeks (9.00-18.7) and 11 weeks (6.14-14.0) after the last dose for Q4W and Q8W, respectively, during pregnancy.

Predicted cord-maternal blood ratio was greater than 1 for cabotegravir, but less than 1 for rilpivirine close to delivery (Table II). Whilst predicted first quartile cord plasma levels were above 4*PA-IC_90_ for cabotegravir, predicted third quartile cord plasma levels were below PA-IC_90_ for rilpivirine (Table II). Likewise, subtherapeutic plasma concentrations of cabotegravir might persist for a considerable period in the mother during lactation (Table III).

**Table 2:** Predicted foetal exposure during the tail-phase pharmacokinetics of LA-CAB/RPV in pregnancy.

| Gestation age | Every 4 weeks dosing |  | Every 8 weeks dosing |  |
| --- | --- | --- | --- | --- |
|  | CAB | RPV | CAB | RPV |
| **Cord plasma concentration (ng/mL)** |  |  |  |  |
| Week 15 | 132 (123-140) | 0.80 (0.75-0.86) | 112 (103-123) | 0.67 (0.63-0.72) |
| Week 27 | 1432 (1342-1564) | 13.7 (13.0-14.8) | 1220 (1105-1357) | 11.5 (10.9-12.4) |
| Week 38 | 837 (786-914) | 11.0 (10.4-11.8) | 715 (648-792) | 9.20 (8.73-9.91) |
| Week 40 | 764 (717-834) | 10.2 (9.7-11.0) | 652 (592-723) | 8.56 (8.12-9.22) |
| **Cord-maternal blood ratio** |  |  |  |  |
| Week 15 | 0.07 | 0.02 | 0.07 | 0.02 |
| Week 27 | 1.69 | 0.57 | 1.69 | 0.57 |
| Week 38 | 1.84 | 0.85 | 1.84 | 0.85 |
| Week 40 | 1.85 | 0.88 | 1.85 | 0.88 |

Concentration data presented as median (interquartile range). Abbreviations: CAB – cabotegravir; RPV – rilpivirine.

**Table 3:** Predicted maternal concentrations of LA-CAB/RPV at delivery and postpartum.

| Maternal plasma<br>concentration (ng/mL) | Every 4 weeks dosing |  | Every 8 weeks dosing |  |
| --- | --- | --- | --- | --- |
|  | CAB | RPV | CAB | RPV |
| At delivery | 415 (386-448) | 11.6 (11.0-12.6) | 353 (318-393) | 9.80 (9.21-10.6) |
| 6 months postpartum | 125 (115-139) | 7.84 (7.30-8.49) | 105 (96.0-116) | 6.82 (6.30-7.42) |

Concentration data presented as median (interquartile range). Abbreviations: CAB – cabotegravir; RPV – rilpivirine.

## Discussion

In this study, an original approach was employed to simulate the tail-phase disposition of LA-CAB/RPV when discontinued early in pregnancy, but after reaching steady-state prior to conception. For the first time, predicted fetal exposure to LA-CAB/RPV during this tail-phase kinetics in the second and third trimesters of pregnancy is described as well as the time to fall below 4*PAIC_90_ for LA-CAB/RPV. Model predictions suggest that both cabotegravir and rilpivirine could remain detectable in the maternal plasma throughout pregnancy after dosing discontinuation early in pregnancy. With lower limits of quantitation of cabotegravir and rilpivirine previously reported as 25 and 1 ng/mL (12, 50), respectively, both drugs are also predicted to remain detectable in the maternal plasma up to 6 months postpartum. However, predicted plasma drug concentrations fell below the 4*PA-IC_90_ level for both drugs in all the virtual maternal population before delivery. In addition, predicted rilpivirine concentration in the cord blood at delivery was below the PA-IC_90_ (12 ng/mL). In contrast, predicted cabotegravir concentration in the cord blood was generally above the PA-IC_90_ (166 ng/mL).

The long tail-phase of LA-CAB/RPV after discontinuation is chiefly influenced by the slow-release nature of the drugs from the muscle depot where it is administered. Model predictions show that the administered drug accumulates within the muscles with each dosing until steady state is reached around 21 months. In addition, the size of the drug depot in the muscle was predicted to reach about 380 and 170% of the maintenance dose for both drugs at steady state with Q4W and Q8W dosing, respectively.

A previous study has reported that cabotegravir remained detectable up to 76 weeks after dosing discontinuation in non-pregnant adults (12). Another study also reported the long tail-phase of LA-CAB/RPV after discontinuation in pregnancy among study participants who discontinued LA-CAB/RPV upon becoming pregnant. Though some of the study participants had not achieved steady state before the discontinuation in pregnancy, both cabotegravir and rilpivirine remained detectable until postpartum in most of the study participants. However, the drug concentrations within the cord blood at delivery was not reported (25).

Ideally, pregnant women taking LA-CAB/RPV for HIV treatment should be immediately switched to a suitable oral regimen upon discontinuation of the LAI option. However, strategies to minimise the unknown risk associated with LA-CAB for PrEP use during pregnancy might need to be reviewed in the light of ongoing long-term exposure after discontinuation. This could also likely apply to lenacapavir designed for once-yearly dosing (51). Women of child-bearing age who begin LAI drugs for PrEP might need to be pre-informed of the risk of fetal drug exposure should they become pregnant as it would be difficult to prevent fetal exposure even when the LAI is discontinued early in pregnancy. Also, residual concentrations of both drugs could persist within the maternal plasma long after the discontinuation where the LAI is discontinued in pregnancy or shortly before pregnancy. Such residual exposures could also result in infant exposure via breastfeeding. As postpartum physiology reverses towards pre-pregnancy states and depot drug release persists, milk drug transfer may contribute to infant drug exposure with limited benefit-risk. However, this was not investigated in this study.

With subtherapeutic concentrations remaining in the maternal plasma for a considerable time after discontinuation, there might also be a potential risk of the development of drug resistance during the tail-phase especially if no new regimen is introduced upon discontinuation by the patient. This might have clinical implications for women on PrEP with LA-CAB if they discontinue the injections early in pregnancy. Our results support considering an oral PrEP coverage strategy during the tail preferably before subtherapeutic concentrations are reached preferably before 27 weeks after the last dose with the Q8W dosing of LA-CAB. However, the optimal choice of PrEP cover and the minimum duration of the PrEP cover would require further study.

A limitation of this study was that the entire drug disposition of the LAI drugs could not be investigated with a single run in a unitary PBPK model. Therefore, separate PBPK models were used to simulate the drug disposition till steady state was achieved as well as the tail-phase kinetics across different stages of pregnancy and postpartum. Thus, discontinuities in the simulation of the drug disposition are likely despite the carryover of the remaining drug in the depot mass. However, the model predictions for the different stages were combined during data analysis. Another limitation of this study was the use of an adult PBPK model to simulate the PK during postpartum. The postpartum is characterised with the rapid reversal of some pregnancy-induced changes which might influence pharmacokinetics also the mammary gland was not represented in the adult model. Thus, milk-transfer of LA-CAB/RPV were not explored in this study.

## Conclusion

The discontinuation of LA-CAB/RPV shortly before or early in pregnancy is likely to result in fetal exposure to the discontinued drug throughout gestation. Potentially, infant exposure via breastfeeding could also occur as residual drug concentrations are likely to remain detectable in maternal plasma postpartum. Women discontinuing LA-CAB for PrEP might need to switch to oral PrEP to reduce the risk of the development of drug resistance. Further studies are required to investigate the necessary duration for an oral PrEP after discontinuing LA-CAB.

## Data Availability

All data produced in the present study are available upon reasonable request to the authors.

## Author contributions

SA and AO conceptualized the study, SA and AO developed the study methodology, SA conducted the study investigation and analysed the data, AO and CW provided the study resources, SA drafted the manuscript, and all authors reviewed and edited the manuscript.

---

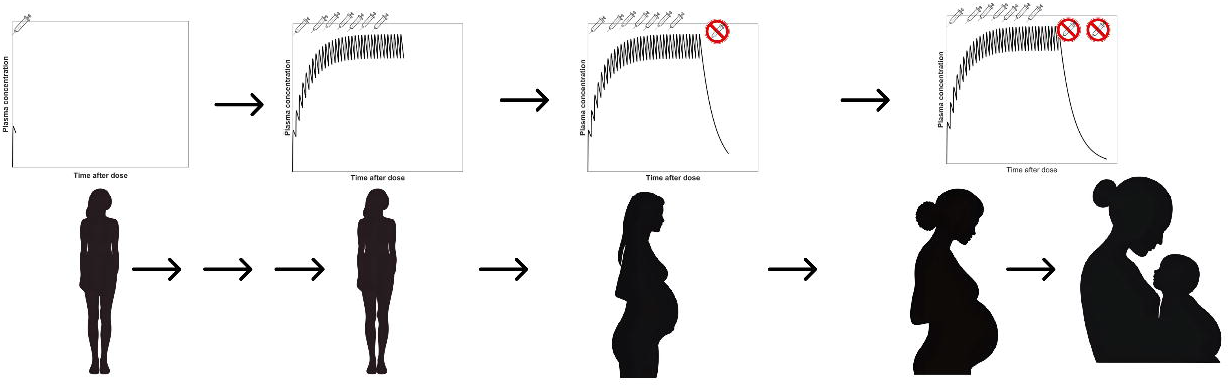

## References

1. UNAIDS. Global HIV & AIDS statistics — Fact sheet 20242025 09 June 2025. Available from: https://www.unaids.org/en/resources/fact-sheet.

2. Trickey A, Sabin CA, Burkholder G, Crane H, d’Arminio Monforte A, Egger M, et al. Life expectancy after 2015 of adults with HIV on long-term antiretroviral therapy in Europe and North America: a collaborative analysis of cohort studies. The Lancet HIV. 2023;10(5):e295–e307.

3. Gudala GR, Padayachee N, Vagiri RV. Beyond antiretroviral treatment: Health-related quality of life of patients receiving antiretroviral treatment at a tertiary hospital in South Africa. Dialogues in Health. 2025;6:100207.

4. Okere NE, Censi V, Machibya C, Costigan K, Katambi P, Martelli G, et al. Beyond viral suppression: Quality of life among stable ART clients in a differentiated service delivery intervention in Tanzania. Qual Life Res. 2022;31(1):159–70.

5. Rosen S, Maskew M, Fox MP, Nyoni C, Mongwenyana C, Malete G, et al. Initiating Antiretroviral Therapy for HIV at a Patient’s First Clinic Visit: The RapIT Randomized Controlled Trial. PLoS Med. 2016;13(5):e1002015.

6. Labhardt ND, Ringera I, Lejone TI, Klimkait T, Muhairwe J, Amstutz A, Glass TR. Effect of Offering Same-Day ART vs Usual Health Facility Referral During Home-Based HIV Testing on Linkage to Care and Viral Suppression Among Adults With HIV in Lesotho: The CASCADE Randomized Clinical Trial. Jama. 2018;319(11):1103–12.

7. Chesney MA. Factors Affecting Adherence to Antiretroviral Therapy. Clinical Infectious Diseases. 2000;30(Supplement_2):S171–S6.

8. Alhassan Y, Twimukye A, Malaba T, Myer L, Waitt C, Lamorde M, et al. ‘I fear my partner will abandon me’: the intersection of late initiation of antenatal care in pregnancy and poor ART adherence among women living with HIV in South Africa and Uganda. BMC Pregnancy Childbirth. 2022;22(1):566.

9. Benítez-Gutiérrez L, Soriano V, Requena S, Arias A, Barreiro P, de Mendoza C. Treatment and prevention of HIV infection with long-acting antiretrovirals. Expert Rev Clin Pharmacol. 2018;11(5):507–17.

10. Martin M, Vanichseni S, Suntharasamai P, Sangkum U, Mock PA, Leethochawalit M, et al. The impact of adherence to preexposure prophylaxis on the risk of HIV infection among people who inject drugs. AIDS. 2015;29(7):819–24.

11. Scarsi KK, Swindells S. The Promise of Improved Adherence With Long-Acting Antiretroviral Therapy: What Are the Data? Journal of the International Association of Providers of AIDS Care (JIAPAC). 2021;20:23259582211009011.

12. Landovitz RJ, Li S, Eron JJ, Jr., Grinsztejn B, Dawood H, Liu AY, et al. Tail-phase safety, tolerability, and pharmacokinetics of long-acting injectable cabotegravir in HIV-uninfected adults: a secondary analysis of the HPTN 077 trial. The Lancet HIV. 2020;7(7):e472–e81.

13. Atoyebi S, Bunglawala F, Cottura N, Grañana-Castillo S, Montanha MC, Olagunju A, et al. Physiologically-based pharmacokinetic modelling of long-acting injectable cabotegravir and rilpivirine in pregnancy. British Journal of Clinical Pharmacology. 2025;91(4):989–1002.

14. Thoueille P, Choong E, Cavassini M, Buclin T, Decosterd LA. Long-acting antiretrovirals: a new era for the management and prevention of HIV infection. J Antimicrob Chemother. 2022;77(2):290–302.

15. Newell M-L. Mechanisms and timing of mother-to-child transmission of HIV-1. AIDS. 1998;12(8):831–7.

16. McGowan JP, Shah SS. Prevention of perinatal HIV transmission during pregnancy. Journal of Antimicrobial Chemotherapy. 2000;46(5):657–68.

17. Cardenas MC, Farnan S, Hamel BL, Mejia Plazas MC, Sintim-Aboagye E, Littlefield DR, et al. Prevention of the Vertical Transmission of HIV; A Recap of the Journey so Far. Viruses. 2023;15(4):849.

18. Fowler MG, Qin M, Fiscus SA, Currier JS, Flynn PM, Chipato T, et al. Benefits and risks of antiretroviral therapy for perinatal HIV prevention. New England Journal of Medicine. 2016;375(18):1726–37.

19. Abduljalil K, Furness P, Johnson TN, Rostami-Hodjegan A, Soltani H. Anatomical, physiological and metabolic changes with gestational age during normal pregnancy: a database for parameters required in physiologically based pharmacokinetic modelling. Clinical Pharmacokinetics. 2012;51(6):365–96.

20. Coppola P, Butler A, Cole S, Kerwash E. Total and Free Blood and Plasma Concentration Changes in Pregnancy for Medicines Highly Bound to Plasma Proteins: Application of Physiologically Based Pharmacokinetic Modelling to Understand the Impact on Efficacy. Pharmaceutics. 2023;15(10):2455.

21. Abduljalil K, Pansari A, Jamei M. Prediction of maternal pharmacokinetics using physiologically based pharmacokinetic models: assessing the impact of the longitudinal changes in the activity of CYP1A2, CYP2D6 and CYP3A4 enzymes during pregnancy. J Pharmacokinet Pharmacodyn. 2020;47(4):361–83.

22. Liu XI, Momper JD, Rakhmanina NY, Green DJ, Burckart GJ, Cressey TR, et al. Prediction of Maternal and Fetal Pharmacokinetics of Dolutegravir and Raltegravir Using Physiologically Based Pharmacokinetic Modeling. Clinical pharmacokinetics. 2020;59(11):1433–50.

23. Atoyebi S, Bunglawala F, Cottura N, Grañana-Castillo S, Montanha MC, Olagunju A, et al. Physiologically-based pharmacokinetic modelling of long-acting injectable cabotegravir and rilpivirine in pregnancy. British journal of clinical pharmacology. 2025;91(4):989–1002.

24. McCormack SA, Best BM. Protecting the fetus against HIV infection: a systematic review of placental transfer of antiretrovirals. Clinical pharmacokinetics. 2014;53(11):989–1004.

25. Patel P, Ford SL, Baker M, Meyer C, Garside L, D’Amico R, et al. Pregnancy outcomes and pharmacokinetics in pregnant women living with HIV exposed to long-acting cabotegravir and rilpivirine in clinical trials. HIV Medicine. 2023;24(5):568–79.

26. Coleman H, Fox J, Chilton D. The risks associated with stopping injectable antiretroviral treatment in women who are trying to conceive: a case series. AIDS. 2022;36(8):1205–6.

27. Jones HM, Maurice D, Kuresh Y, R. GJ, J. AN, L. HT, et al. Application of PBPK modelling in drug discovery and development at Pfizer. Xenobiotica. 2012;42(1):94–106.

28. Montanha MC, Cottura N, Booth M, Hodge D, Bunglawala F, Kinvig H, et al. PBPK Modelling of Dexamethasone in Patients With COVID-19 and Liver Disease. Front Pharmacol. 2022;13:814134.

29. Montanha MC, Fabrega F, Howarth A, Cottura N, Kinvig H, Bunglawala F, et al. Predicting Drug–Drug Interactions between Rifampicin and Ritonavir-Boosted Atazanavir Using PBPK Modelling. Clinical Pharmacokinetics. 2022;61(3):375–86.

30. Cottura N, Kinvig H, Grañana-Castillo S, Wood A, Siccardi M. Drug-Drug Interactions in People Living With HIV at Risk of Hepatic and Renal Impairment: Current Status and Future Perspectives. J Clin Pharmacol. 2022;62(7):835–46.

31. Sun Z, Zhao N, Xie R, Jia B, Xu J, Luo L, et al. Physiologically-based pharmacokinetic modeling predicts the drug interaction potential of GLS4 in co-administered with ritonavir. CPT: Pharmacometrics & Systems Pharmacology. 2024;13(9):1503–12.

32. Chen S, Shen C, Tian Y, Peng Y, Hu J, Xie H, Yin P. Physiologically based pharmacokinetic modeling and simulation of topiramate in populations with renal and hepatic impairment and considerations for drug–drug interactions. CPT: Pharmacometrics & Systems Pharmacology. 2024;n/a(n/a).

33. Atoyebi S, Montanha MC, Nakijoba R, Orrell C, Mugerwa H, Siccardi M, et al. Physiologically based pharmacokinetic modeling of drug–drug interactions between ritonavir-boosted atazanavir and rifampicin in pregnancy. CPT: Pharmacometrics & Systems Pharmacology. 2024;n/a(n/a).

34. Bunglawala F, Rajoli RKR, Mirochnick M, Owen A, Siccardi M. Prediction of dolutegravir pharmacokinetics and dose optimization in neonates via physiologically based pharmacokinetic (PBPK) modelling. Journal of Antimicrobial Chemotherapy. 2020;75(3):640–7.

35. van Hoogdalem MW, Tanaka R, Abduljalil K, Johnson TN, Wexelblatt SL, Akinbi HT, et al. Forecasting Fetal Buprenorphine Exposure through Maternal-Fetal Physiologically Based Pharmacokinetic Modeling. Pharmaceutics. 2024;16(3).

36. Shenkoya B, Gopalakrishnan M, Eke AC. Physiologically based pharmacokinetic modeling of long-acting extended-release naltrexone in pregnant women with opioid use disorder. CPT Pharmacometrics Syst Pharmacol. 2024;13(11):1939–52.

37. Gill KL, Jones HM. Opportunities and Challenges for PBPK Model of mAbs in Paediatrics and Pregnancy. The AAPS journal. 2022;24(4):72.

38. Bediako-Kakari P, Monyo M, Atoyebi S, Olagunju A. Comparative modelling of foetal exposure to maternal long-acting injectable versus oral daily antipsychotics. npj Women’s Health. 2025;3(1):31.

39. Spreen W, Williams P, Margolis D, Ford SL, Crauwels H, Lou Y, et al. Pharmacokinetics, safety, and tolerability with repeat doses of GSK1265744 and rilpivirine (TMC278) long-acting nanosuspensions in healthy adults. J Acquir Immune Defic Syndr. 2014;67(5):487–92.

40. Ford SL, Sutton K, Lou Y, Zhang Z, Tenorio A, Trezza C, et al. Effect of Rifampin on the Single-Dose Pharmacokinetics of Oral Cabotegravir in Healthy Subjects. Antimicrob Agents Chemother. 2017;61(10).

41. Center for Drug Evaluation and Research. Clinical Pharmacology and Biopharmaceutics Review(s). Addendum to Ondqa Biopharmaceutics review. 2011.

42. Crauwels H, Vingerhoets J, Ryan R, Witek J, Anderson D. Pharmacokinetic Parameters of Once-Daily Rilpivirine following Administration of Efavirenz in Healthy Subjects. Antiviral Therapy. 2012;17(3):439–46.

43. Rajoli RKR, Curley P, Chiong J, Back D, Flexner C, Owen A, Siccardi M. Predicting Drug-Drug Interactions Between Rifampicin and Long-Acting Cabotegravir and Rilpivirine Using Physiologically Based Pharmacokinetic Modeling. J Infect Dis. 2019;219(11):1735–42.

44. Watts DH, Stek A, Best BM, Wang J, Capparelli EV, Cressey TR, et al. Raltegravir pharmacokinetics during pregnancy. J Acquir Immune Defic Syndr. 2014;67(4):375–81.

45. Osiyemi O, Yasin S, Zorrilla C, Bicer C, Hillewaert V, Brown K, Crauwels HM. Pharmacokinetics, Antiviral Activity, and Safety of Rilpivirine in Pregnant Women with HIV-1 Infection: Results of a Phase 3b, Multicenter, Open-Label Study. Infectious Diseases and Therapy. 2018;7(1):147–59.

46. Schalkwijk S, Colbers A, Konopnicki D, Gingelmaier A, Lambert J, van der Ende M, et al. Lowered Rilpivirine Exposure During the Third Trimester of Pregnancy in Human Immunodeficiency Virus Type 1–Infected Women. Clinical Infectious Diseases. 2017;65(8):1335–41.

47. Ford SL, Gould E, Chen S, Margolis D, Spreen W, Crauwels H, Piscitelli S. Lack of pharmacokinetic interaction between rilpivirine and integrase inhibitors dolutegravir and GSK1265744. Antimicrobial agents and chemotherapy. 2013;57(11):5472–7.

48. ViiV Healthcare. CABENUVA (cabotegravir extended-release injectable suspension; rilpivirine extended-release injectable suspension). US Prescribing Information. 2022.

49. European Medicines Agency. Apretude (cabotegravir) 2023 [updated 20/09/2023. Available from: https://www.ema.europa.eu/en/medicines/human/EPAR/apretude.

50. Neyens M, Crauwels HM, Perez-Ruixo JJ, Rossenu S. Population pharmacokinetics of the rilpivirine long-acting formulation after intramuscular dosing in healthy subjects and people living with HIV. Journal of Antimicrobial Chemotherapy. 2021;76(12):3255–62.

51. Jogiraju V, Pawar P, Yager J, Ling J, Shen G, Chiu A, et al. Pharmacokinetics and safety of once-yearly lenacapavir: a phase 1, open-label study. The Lancet. 2025;405(10485):1147–54.

